# Missed Golden Hour? Proportion and Factors Associated with Timely Specialist Review of Very High-Risk Obstetric Mothers in Eastern Uganda: A Retrospective Study

**DOI:** 10.64898/2026.08.18.26360660

**Authors:** Ronald Tweheyo, Shamim Nabidda, Proscovia Auma, Baifa Alwenyo, Jude Mulowooza, Andrew Twineamasiko, Mildred Irene Neumbe, Jonathan Babuya, Frank Kayemba, Simon Odoch, Dan Kibuule, Paul Waako, Stephen Obbo, Enid Kawala Kagoya

## Abstract

**Background:** Timely review of very high-risk mothers by an obstetrician within one hour of admission is very important in enabling fast decision making for emergency intervention. Any delays in review of high-risk obstetric patient such as hypertensive disorders, obstructed labour or haemorrhages increase maternal morbidity and mortality. Globally, more than 260,000 mothers die from pregnancy related causes with sub-Saharan Africa being contributing 70%. To reduce this mortality, the ministry of health of Uganda encourages urgent assessment of all high-risk pregnant mothers. This study assessed the proportion and factors associated with specialist review within one hour of very high-risk obstetric mothers at Mbale Regional Referral Hospital.

**Methods:** A retrospective quantitative study was conducted from June to October 2025 at a tertiary Hospital in Easter Uganda. Systematic sampling was used to select files of mothers triaged as very high (red category). The minimum calculated sample size was 427, but 454 eligible files were analysed to improve precision. Social demographics obstetric characteristics and timing of specialist review ere extracted. Data were entered into excel and analysed using STATA. Descriptive statistics summarized proportions and modified Poisson regression identified factors associated with timely review at 95% CI and p<005.

**Results:** The proportion of very high-risk mothers reviewed within one hour was 33.9 % (95% CI:29.7%-38.4%). In multivariable analysis, foetal heart monitoring conducted once was independently associated with lower likelihood of timely review (aPR=0.575,95% CI:0.334-0.988; p=0.045). No other variables showed significant association.

**Conclusion:** Only one-third of very high-risk mothers received specialist review within one hour below national recommendations. Strengthening obstetric triage and specialist availability is essential to improving emergency obstetric care

## Introduction

Timely review of very high-risk mothers refers to assessment by a senior obstetric clinician within one hour of hospital admission to facilitate rapid decision-making necessary to initiate appropriate emergency care (1, 2). This early obstetric evaluation is very critical in obstetric emergency care, particularly for mothers presenting with life-threatening conditions such as hypertensive disorders, obstructed labour, haemorrhage, and other obstetric emergencies (3). Delay of obstetric emergency interventions for very high-risk obstetric mothers results in increased risk of complications and maternal morbidity and mortality (4, 5)

Globally, maternal and perinatal morbidity remain high, with about 260 000 women dying during and following pregnancy and childbirth (6). Low- and middle-income countries have the highest maternal mortality globally at 92% (6). In sub-Saharan Africa, maternal mortality accounts for about two-thirds of global maternal deaths, highlighting persistent challenges in accessing timely and quality obstetric care (7, 8) The majority of these deaths are usually preventable, but because of a lack of timely response to emergencies, deaths always become inevitable (9, 10). In Uganda, 189 out of 100,000 women die every year(6). However, there is little documentation about the exact number that dies in Uganda due to delayed review by specialists.

According to the Ministry of Health in Uganda, all high-risk mothers(100%) presenting with obstetric emergencies or high-risk conditions should be assessed and managed urgently with appropriate senior clinical input to prevent maternal morbidity and (11).This recommendation shows national commitment to improving maternal outcomes throughout obstetric care (11). However, the above has been had to achieve due to many health faculties having senior clinicians (obstetricians) facing multiple competing interests and responsibilities such as covering many wards, operating theatres and referral duties across facilities (12). This usually limits their availability for immediate assessment of high-risk mothers (3). The shortage of obstetric specialists, high patient volumes, and inadequate support staff also contribute to delays in specialist review (3).

These systematic and resource-related constraints can result in delayed decision-making and delayed interventions. Understanding these barriers is essential to designing interventions that ensure timely senior clinical input for high-risk obstetric cases (3). This study was aimed at assessing the number of high-risk mothers seen by a specialist within one hour of hospital admission.

### Study Design and Setting

This study employed a retrospective quantitative study approach. This method was used to determine the prevalence with specialist review within one hour among very high-risk obstetric mothers at Mbale Regional Hospital, Eastern Uganda. This study was done by reviewing files of mothers who had been admitted to Mbale Regional Referral Hospital from June 2025 to October 2025. The number of mothers triaged each week had their files monitored on weather a specialist had seen them within one hour of admission or not. Social demographics, together with obstetric characteristics, were also extracted from the files. The results were integrated into the discussion section. The study was done at Mbale Regional Referral Hospital (MRRH), a public tertiary hospital located in Eastern Uganda. This Hospital Usually admits about 735 to 980 per week (13). Most of these mothers are referred from peripheral facilities due to anticipated complications; some are self-referred patients, while others are non-referral admissions.

### Study Population and Sampling

This study included only files of mothers who had been admitted to Mbale Regional Referral Hospital from June 2025 to October 2025. Some were referred mothers, while others were non-referred. The sampling method employed was a systematic sampling of only mothers triaged as high-risk mothers (red) admitted at Mbale Regional Referral Hospital from June 2025 to October 2025. In this study, 1,087 patient files were selected and used for the study.

### Sample size calculation

In this study, sample size was calculated using ( Leslie kish Formulan1965) (14), Where,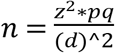 Hence, the figure is represented as follows; n=desired sample size. Z=standard normal deviation, which is equal to 1.96, corresponding to a 95% confidence interval p= 0.52 and q= (1-0.52) = 0.48, and d=the standard error, d = 0.05.

The sample size was also based on the study that was carried out in Malawi, where 52% of women died at a healthcare facility 52.1% due to delay in deciding to seek care, reaching the healthcare facility; and receiving care at the healthcare facility (15). This study was used because it was the one closely related to the study we did (15).

Substituting into the formula is as follows.

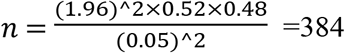patient files. Adjusting for a 10% non-response rate (384÷0.9) =427 files as the minimum sample size. In this study, we included more than the minimum required sample size of (454) files more than the minimum sample size of 427 by 27 files, to improve the precision of estimates and statistical power, ensuring adequate data even if some responses are incomplete (16)

### Study Variables

The independent variables were triage, presence or absence of a sticker, colour of sticker, being assessed with triage form, monitoring and taking of foetal heart rate, time of admission (day or night) as well as day of admission (weekday or weekend.

The primary dependent variable (outcome variable) was a high-risk mother (triaged as red) being reviewed by a specialist within one hour. This was measured by reviewing the time when the patient was admitted and when the patient’s file had documentation of being reviewed by a specialist within one hour.

### Data collection method

The data were collected using a retrospective review of patient files. The data was collected in two steps. In the first step, relevant patient files were identified, and later data were collected by research assistants, who were trained students and nurses, and the supervisor was a specialist obstetrician. This data was collected in 1^st^ June to 20^th^ June2026

### Data analysis

All data were checked appropriately after collection to ensure authenticity and completeness. Data was first entered into Microsoft Excel and later imported into STATA for analysis. The descriptive statistics that were used were frequencies and percentages for categorical data and means and medians for continuous data. Logistic regression was carried out to determine the factors associated with a high-risk mother being reviewed by a specialist within one hour at Mbale Regional Referral Hospital. Bivariate and multivariate analyses were also used to control for confounders. The significance of the investigated variables was determined when the p-values were less than 0.05 at a 95% confidence level.

### Ethical approval

Ethical approval was obtained from the Busitema University Research Ethics Committee, reference number BUFHS-2026-691, and administrative clearance was obtained from the Mbale Regional Referral Hospital. The study was done according to the Declaration of Helsinki. As this was a retrospective review of routinely collected medical records, the Research Ethics Committee waived the requirement for informed consent. Patient identifiers were not collected, and all data were anonymised to ensure confidentiality. Patient codes were used instead of their real names to protect participants’ identities.

## Result

Most women had no triage documented (76.9%, n=349). A foetal heart rate was taken on admission for 91.0% (n=413), and foetal heart monitoring was documented for 94.9% (n=431). Among women with the monitoring frequency recorded (n=452), monitoring was most commonly done three times (49.8%, n=225) or twice (41.6%, n=188).

By diagnosis (n=454), uncomplicated labour was the largest category (35.2%, n=160), followed by other obstetric emergencies (30.6%, n=139). Admissions occurred more often on weekends (75.5%, n=335) than on weekdays, and slightly more often at night (51.8%, n=235) than during the day (44.9%, n=204). Most babies were recorded as alive (99.8%, n=416), and most mothers delivered (91.9%, n=417) (**Table 1**).

**Table 1.**
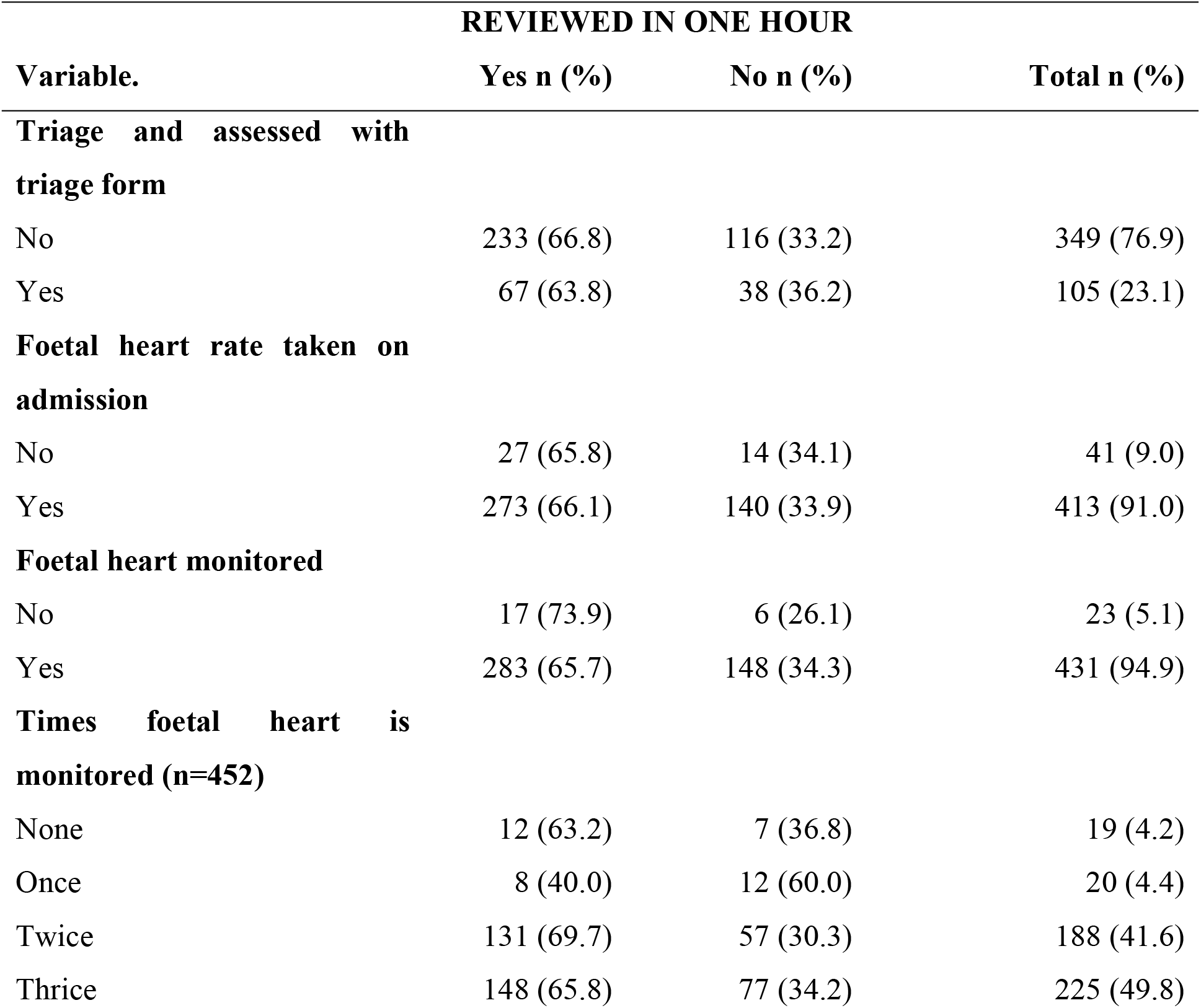
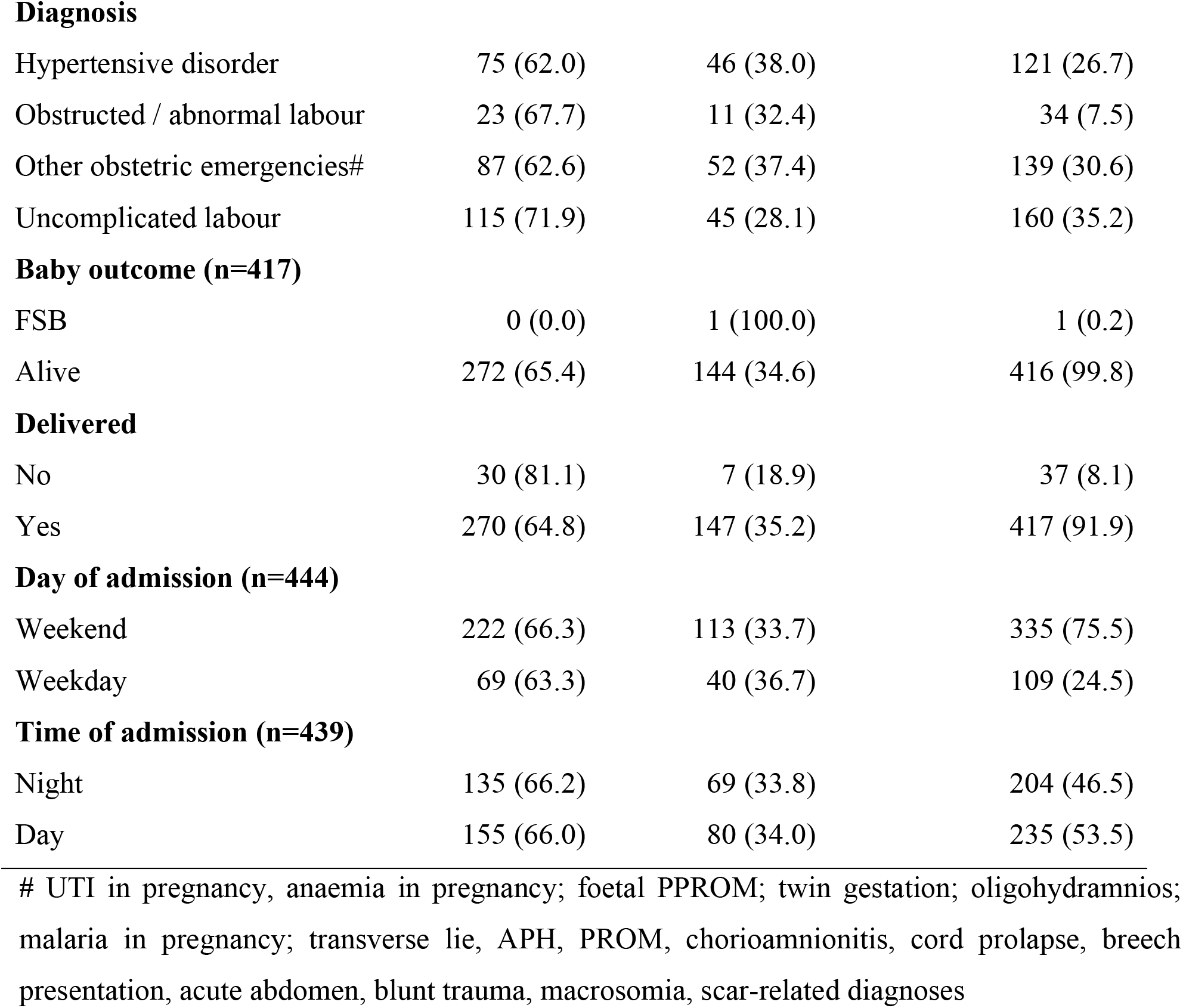
Triage, clinical and admission-timing factors associated with specialist review within one hour among 454 mothers at MRRH.

| Variable. | REVIEWED IN ONE HOUR |  |  |
| --- | --- | --- | --- |
|  | Yes n (%) | No n (%) | Total n (%) |
| **Triage and assessed with triage form** |  |  |  |
| No | 233 (66.8) | 116 (33.2) | 349 (76.9) |
| Yes | 67 (63.8) | 38 (36.2) | 105 (23.1) |
| **Foetal heart rate taken on admission** |  |  |  |
| No | 27 (65.8) | 14 (34.1) | 41 (9.0) |
| Yes | 273 (66.1) | 140 (33.9) | 413 (91.0) |
| **Foetal heart monitored** |  |  |  |
| No | 17 (73.9) | 6 (26.1) | 23 (5.1) |
| Yes | 283 (65.7) | 148 (34.3) | 431 (94.9) |
| **Times foetal heart is monitored (n=452)** |  |  |  |
| None | 12 (63.2) | 7 (36.8) | 19 (4.2) |
| Once | 8 (40.0) | 12 (60.0) | 20 (4.4) |
| Twice | 131 (69.7) | 57 (30.3) | 188 (41.6) |
| Thrice | 148 (65.8) | 77 (34.2) | 225 (49.8) |
| **Diagnosis** |  |  |  |
| Hypertensive disorder | 75 (62.0) | 46 (38.0) | 121 (26.7) |
| Obstructed / abnormal labour | 23 (67.7) | 11 (32.4) | 34 (7.5) |
| Other obstetric emergencies# | 87 (62.6) | 52 (37.4) | 139 (30.6) |
| Uncomplicated labour | 115 (71.9) | 45 (28.1) | 160 (35.2) |
| **Baby outcome (n=417)** |  |  |  |
| FSB | 0 (0.0) | 1 (100.0) | 1 (0.2) |
| Alive | 272 (65.4) | 144 (34.6) | 416 (99.8) |
| **Delivered** |  |  |  |
| No | 30 (81.1) | 7 (18.9) | 37 (8.1) |
| Yes | 270 (64.8) | 147 (35.2) | 417 (91.9) |
| **Day of admission (n=444)** |  |  |  |
| Weekend | 222 (66.3) | 113 (33.7) | 335 (75.5) |
| Weekday | 69 (63.3) | 40 (36.7) | 109 (24.5) |
| **Time of admission (n=439)** |  |  |  |
| Night | 135 (66.2) | 69 (33.8) | 204 (46.5) |
| Day | 155 (66.0) | 80 (34.0) | 235 (53.5) |

### The proportion of very high-risk obstetric patients reviewed by a specialist within one hour of admission

Proportion of very high-risk obstetric patients reviewed by a specialist within one hour of admission was 33.9% (95% confidence interval (CI): 29.7%-38.4%) (Figure 1).

**Figure 1.**
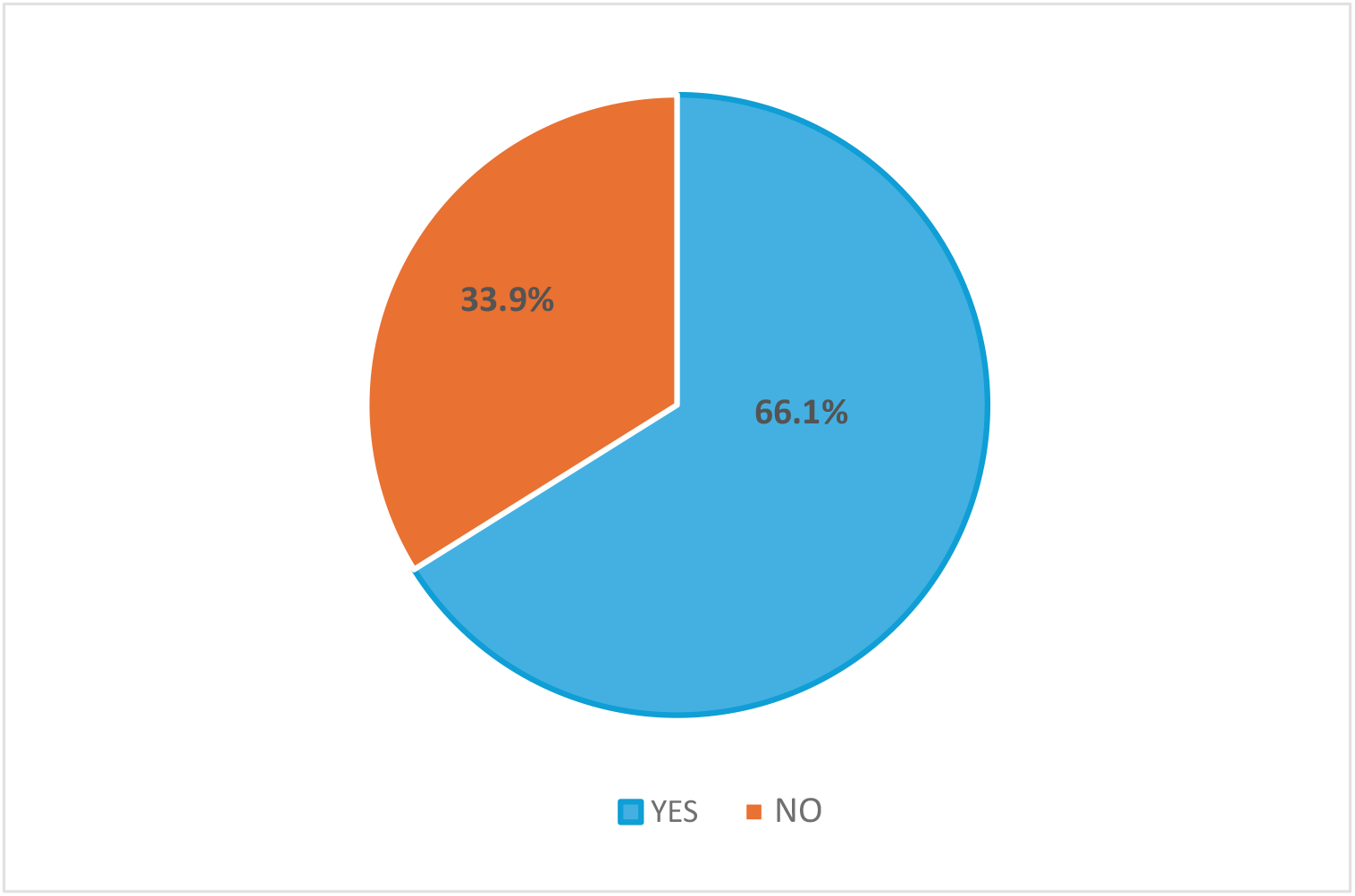
Proportion of very high-risk obstetric patients reviewed by a specialist within one hour of admission.

### Bivariate analysis and multivariate analysis of factors associated with review of very high-risk obstetric mothers by a specialist within one hour at MRRH

At bivariate analysis, **diagnosis** (hypertensive disorders: cPR = 0.862, 95% CI: 0.727-1.022; p = 0.088), (Other obstetric emergencies: cPR = 0.871, 95% CI: 0.741-1.023; p = 0.093) and times foetal heart was monitored (once: cPR = 0.574, 95% CI: 0.333-0.991; p = 0.046) met the p<0.20 criterion and were considered for multivariable analysis **(Table 2)**. At multivariable analysis, times foetal heart was monitored (once: aPR = 0.575, 95% CI: 0.334-0.988; p = 0.045) was **associated with timely review by specialist** since it had p-value <0.05 **(Table 2)**.

**Table 2.**
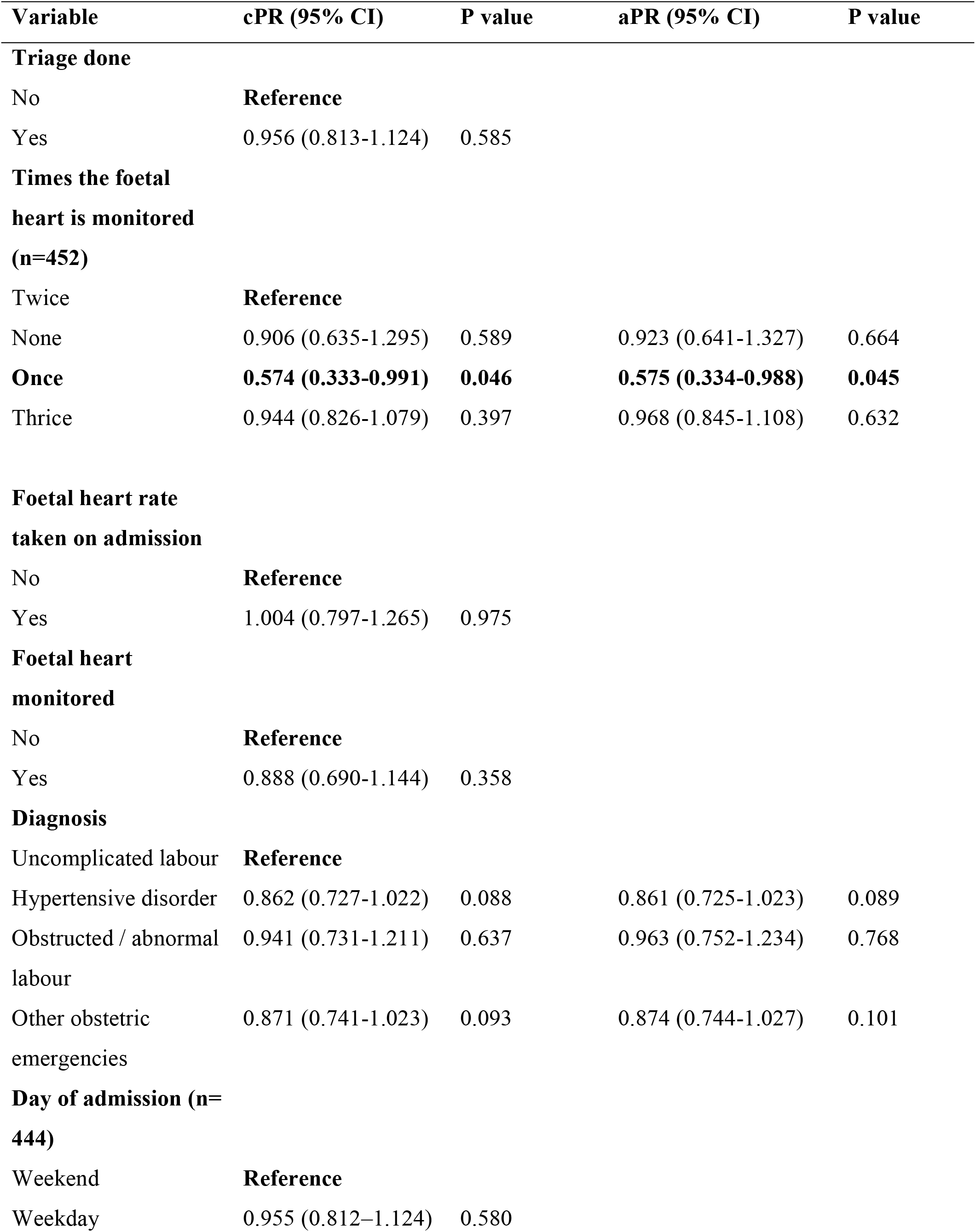
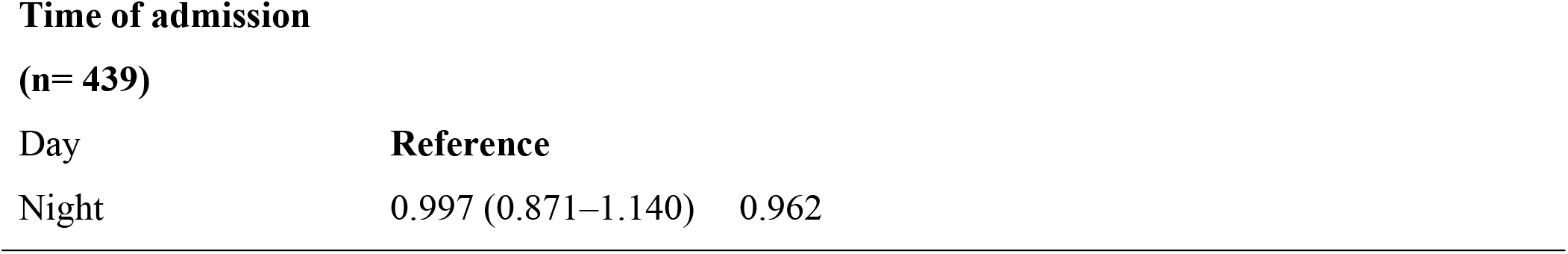
Bivariate analysis and multivariate analysis of factors associated with review of very high-risk obstetric mothers by a specialist within one hour at MRRH.

## Discussion

This study assessed the proportion of very high-risk obstetric mothers reviewed by a specialist within one hour of admission at Mbale Regional Referral Hospital. The study also examined factors associated with timely review. The findings showed that only 33.9% of very high-risk mothers were reviewed by a specialist within one hour, which is below national recommendations for high-risk obstetric patients receiving urgent senior clinical assessment. In the multivariable analysis, foetal heart monitoring conducted once was associated with a lower likelihood of timely specialist review as compared to those that had fetal heart monitoring done twice. Other factors such as triage documentation, diagnosis, day of admission, and time of admission were not significantly associated with timely review. Some of the findings obtained in our study were similar or different to those from other studies.

In this study, we found out that 33.9% (CI: 29.7%-38.4%) of the participants had been reviewed by a specialist within one hour of admission. These findings are similar to the findings of a study carried out in Nigeria, where 38.9% of the mothers in a tertiary hospital had been reviewed within one hour (17). These findings have also been obtained from a study carried out in a tertiary hospital, just like in our study (17). In our study, foetal heart monitoring conducted twice was associated with a higher likelihood of timely specialist review as compared to those that had fetal heart monitoring done once. These findings are different from the findings of a scoping review, where staff and space shortages, burnout and fatigue, inadequate knowledge of health professionals, inadequate supplies, poor communication, and absence of responsibility and commitment resulted in unsuccessful implementation of obstetric triage (18). In this study, we also had limited documentation of mothers during triage. This is similar to studies carried out in other low-resource settings where triage systems might be inconsistently applied with incomplete documentation due to staff shortages, heavy workload, or inadequate training(19).Much as our study findings were similar or different from other studies carried out in other countries, our study had some strengths and weaknesses.

The study’s strengths were on provision of useful evidence on the timeliness of specialist review among very high-risk obstetric mothers in a major referral hospital in eastern Uganda. This may help policymakers review the staffing levels of specialists necessary to achieve a 100% review of very high-risk obstetric mothers within one hour at tertiary hospitals in the country.

The study is limited by its reliance on retrospective review of patient files, which entirely had to be considered if the clinical documentation was accurate and complete. Due to this methodology, some events or timings may not have been fully recorded. The study was conducted in a single tertiary hospital, which may limit the generalizability of the findings to other facilities. Despite these limitations, we interpreted our findings in many different ways.

In this study, mothers who were reviewed by a specialist within one hour were only 33.9%. This is a low proportion as compared to the expected 100% of all mothers triaged as red (very high risk). This may reflect challenges commonly faced in sub-Saharan Africa, where initial obstetric assessment of patients is often conducted by midwives, intern doctors, or medical officers, while obstetric specialists are consulted when complications are identified. This approach allows maternity services to function despite shortages of specialists. However, it delays senior clinical involvement at times of admission of very high-risk mothers, especially when their services are needed most. Rushing to quickly judge the specialists would not be of great benefit, as there is a problem of limited specialists, particularly when facilities experience high patient volumes or multiple emergencies simultaneously (20). The act of other health workers, such as intern doctors, midwives, and medical officers, and not necessarily specialists, reviewing patients is usually done for the best interest of the patient. According to the findings from this study and other studies across Africa, delays like this are associated with severe maternal and neonatal outcomes such as eclampsia, pre-eclampsia, severe postpartum haemorrhage, fetal distress, or even death at birth (21, 22). Previous studies from Uganda and other African settings also reported that facility-level delays, including delays in assessment and referral decisions, contribute substantially to poor maternal outcomes(23). Having a few patients being seen by specialists within one hour had many possible causes, one of which could have been poor documentation or limited frequency of fetal heart rate measurements.

In this study, foetal heart monitoring conducted twice was significantly associated with a higher likelihood of timely specialist review as compared to those that had fetal heart monitoring done once. This was because conducting fetal heart rate twice increased the chances of identifying a serious health concern that could have prompted the health worker of lower cadre to seek special specialist consultation amid the tight schedule. Taking heart rate twice could also mean that such patients were referrals from lower health facilities whose referral note required a review by a specialist as soon as possible. In this study, most mothers also had limited documentation in triage, suggesting gaps in the implementation or documentation of obstetric triage systems. This showed that there was ineffective triage in this facility, which possibly also made it hard to quickly identify women with life-threatening conditions and prioritize them for immediate care. This problem can be linked to limited staffing levels, where sufficient documentation amidst tight schedules becomes impossible at the expense of saving lives.

This study informs the importance of improving hospital specialists’ staffing levels to improve service delivery. This will also reduce the mortality rates related to delayed review of specialists within one hour at tertiary hospitals like Mbale Regional Referral Hospital. In order to have good clinical outcomes, pregnant mothers’ fetal heart rate should always be taken at least twice within one hour, as it helps identify a worsening medical condition requiring quick specialist review.

This could also help reduce maternal mortality, particularly in areas with few specialists amidst many sudden, unexpected obstetric emergencies at triage, such as Eastern Uganda and other parts of sub-Saharan Africa. Evidence showed that structured obstetric triage systems improve the timeliness of clinical assessment and reduce waiting times for emergency obstetric care(23, 24). For a long time, shortages of obstetric specialists have remained a major challenge in many African countries, contributing to gaps in access to timely specialist care. Previous studies have shown that staffing patterns, workload, and referral processes may affect the timeliness of emergency obstetric care in hospital settings(19, 23).In many referral hospitals, specialists supervise multiple wards, theatres, and emergency units simultaneously, which may limit their availability for immediate assessment of all incoming obstetric emergencies. Therefore, in the presence of sufficient specialist hospital staffing, good documentation, and vital monitoring such as fetal heart rate, waiting times for obstetric care can be reduced, improving overall clinical outcomes.

## Conclusions

The findings of this study had several implications for improving emergency obstetric care. Strengthening obstetric triage systems, improving documentation practices, and establishing clear protocols for rapid escalation of high-risk cases may help improve the timeliness of specialist review. In addition, increasing the availability of trained obstetric specialists and strengthening collaboration between midwives, medical officers, and specialists may improve the responsiveness and quality of maternity care in referral hospitals.

Overall, the findings highlight important gaps in the timely review of high-risk obstetric mothers by specialists. Improving obstetric triage systems, strengthening documentation practices, and increasing specialist availability may help enhance the quality and responsiveness of emergency obstetric care in tertiary health facilities.

## Availability of data and materials

All relevant data are within the paper and its supporting information files.

## Consent to participate

The requirement for informed consent was waived by the Busitema University Research Ethics Committee (reference number BUFHS-2026-691), and administrative clearance was obtained from the Mbale Regional Referral Hospital. Because this study involved retrospective review of routinely collected patient records, no direct contact with participants occurred, and all data were anonymised before analysis.

## Author’s statement

EKK (Enid Kawala Kagoya), TR (Ronald Tweheyo), and EKK (Enid Kawala Kagoya) conceptualised the study and wrote the proposal; TR, JB (Jonathan Babuya), EKK, SN (Shamin Nabidda), and RT analysed the data and drafted the manuscript; EKK, SN, TR, JB, PA (Proscovia Auma), FK (Frank Kayemba), and BA (Baifa Arwenyo) reviewed the manuscript. TR, EKK, JM (Jude Mulowooza), AT (Andrew Twineamatsiko), SO (Stephen Obbo), and IMN (Irene Milred Neumbe) provided the overall oversight in the design and implementation of the study. TR, PW (Paul Waako), DK (Dan Kibuule), and SO (Simon Odoch) reviewed the final manuscript before submission. All authors meet the criteria for author contributions and have read and approved the manuscript.

## Acknowledgements

We appreciate the participants who agreed to take part in this study.

